# Preserving musculoskeletal health through resistance training in individuals undergoing Glucagon-like Peptide-1 Receptor Agonist Therapy: a controlled interrupted time-series analysis (Stage 1 Registered Report)

**DOI:** 10.1101/2025.06.24.25330195

**Authors:** James Steele, Myles N Moore, Pramuk Mahanma, David Scott, Robin M. Daly

## Abstract

Glucagon-like peptide-1 receptor agonists (GLP-1-RAs) are increasingly prescribed for weight loss and cardiometabolic health but have been evidenced to lead to loss of lean soft tissue mass. Resistance training (RT) is known to result in increase muscle mass, and indeed preserve muscle mass during weight loss through more traditional weight loss approaches such as energy restriction and bariatric surgery. Yet, its effectiveness in counteracting GLP-1-RA–associated lean soft tissue mass loss remains unclear. This Stage 1 Registered Report outlines a quasi-experimental, retrospective, controlled interrupted time-series analysis using existing data from Kieser Australia members undergoing standardized RT intervention. Participants identified by internal survey to have also been on, or are currently on, GLP-1-RA therapy will be propensity score–matched to controls not receiving the drug. The primary outcome is fat free mass (via bioelectrical impedance analysis), serving as a proxy for lean soft tissue mass. We hypothesize that the effect of RT over time will be non-inferior to the effect of GLP-1-RA, indicating mitigation of lean mass loss. Secondary exploratory outcomes include effects on muscle strength. Simulations regarding estimates of GLP-1-RA treatment use in the Kieser Australia membership suggest we will be able to obtain a sample size of 37 [interquartile range: 8] matched participants per group and that, using additive estimates of the effects of RT and GLP-1-RA treatments from prior meta-analyses suggest adequate we will achieved at least 80% power at an alpha of 0.05 for testing non-inferiority with this sample size. Results will inform clinical strategies to preserve musculoskeletal health during pharmacological weight loss interventions.

## Introduction

Obesity is associated with an increased risk for type 2 diabetes mellitus (T2DM), cardiovascular disease, cancer and all-cause mortality (Guh et al., 2009). Traditional treatments for obesity have included lifestyle or surgical-based interventions. However, since the global rate of people with obesity continues to grow (NCD Risk Factor Collaboration (NCD-RisC), 2016) additional treatment strategies that are alternative to traditional methods are necessary to help with the growing burden of this disease.

The prescription of Glucagon-like peptide-1 receptor agonist (GLP-1-RA) treatment has grown between 2014 and 2022 as an effective approach to decrease total body weight and better manage T2DM (Lin et al., 2023). Prescription with GLP-1-RA has also increased in people with cardiovascular conditions without T2DM, likely because there is evidence that this treatment can reduce the risk of cardiovascular events (Lin et al., 2023). Indeed, there is growing evidence for a broad range of health outcomes from these treatments (Olukorode et al., 2024). But despite these health benefits, concerns have been raised (Linge et al., 2024; Prado et al., 2024) that part of the weight loss with GLP-1-RA treatment is due to a decline in lean soft tissue mass (i.e., muscle and organ mass). For example, two recent systematic review and meta-analyses have reported that ∼20-30% of total weight lost during GLP-1-RA treatment was from lean mass (i.e., muscle, bone, and organ mass) with suggestion that this primarily comes from lean soft tissue mass, though a lack of evidence on bone mass specifically (Beavers et al., 2025; Karakasis et al., 2025). This suggests that strategies are needed to preserve the lean soft tissue mass lost during GL-P-1RA treatment.

Progressive resistance training (RT), even at a relatively low training volume, can preserve age-related muscle mass in adults (Benito et al., 2020; Radaelli et al., 2025) and is recommended to preserve muscle mass during traditional weight loss interventions, such as energy restriction through diet (Lopez et al., 2022; Murphy & Koehler, 2022) or bariatric surgery (Morales-Marroquin et al., 2020). Despite this evidence, there are few studies to our knowledge that have examined the effects of RT during GLP-1-RA treatment and whether it may mitigate the loss in lean soft tissue mass that occurs during GLP-1R-A treatment. One study of liraglutide (Jensen et al., 2024) found that lean body mass was maintained in a group undergoing concomitant treatment and exercise intervention; yet the exercise alone group saw increased lean body mass and the liraglutide alone also saw maintenance, so it is unclear that lean body mass loss was present to be mitigated anyway. Given this lack of evidence, additional research is needed to understand whether the effects of RT on muscle mass are sufficiently potent to counteract the GLP-1-RA effects on lean soft tissue loss.

We are fortunate to have access to a potential observational dataset to examine this at Kieser Australia given we have a large membership (∼24,000) of patients who undergo standardized RT intervention, may of which are likely also concomitantly on GLP-1-RA treatment. We also take for many of these patients standardised measurements of body composition over time using bioelectrical impedance analysis (BIA) including fat free mass. We are therefore proposing to complete a retrospective interrupted time-series with control to investigate the effects of GLP-1-RA treatments broadly during participation in RT upon fat free mass as an indicator of lean soft tissue mass^1^.

**Table 1.** Peer Community In registered report design table.

| PRIMARY OUTCOME AND HYPOTHESIS TEST |  |  |  |  |  |  |
| --- | --- | --- | --- | --- | --- | --- |
| Question | Hypothesis | Sampling plan | Analysis plan | Rationale for deciding the sensitivity | Interpretation given different outcomes | Theory that could be shown wrong by the outcome |
| Can resistance training (RT) mitigate lean soft tissue mass loss during Glucagon-like Peptide-1 Receptor Agonist (GLP-1-RA) treatment? | We hypothesize that RT will mitigate lean soft tissue, inferred through fat free mass measured by bioelectrical impedance analysis (BIA), while adults are on GLP-1-RA treatment.<br>Specifically, based on previous meta-analysis of both the magnitude of effects of RT and GLP-1-RA on lean soft tissue mass and an assumption of additive effects (supported by similarly additive effects upon lean soft tissue mass from RT and energy restriction), we hypothesise that the coefficient for the effect of RT over time (fixed effect estimate of time) will be non-inferior to the coefficient for the effect of GLP-1-RA (fixed effect estimate of period:intervention:time) over time from a controlled | We will survey the membership of Kieser Australia via our internal surveying system to identify by self-report members who are currently, or have been previously, on GLP-1-RA treatment. Members responding as having been on the GLP-1-RA treatment will then have their data queried in our database to identify those with fat free mass measurements from BIA. We will then identify via nearest propensity score matching a control group of Kieser members who have not reported having been and also have fat free mass measurements recorded. Intervention and controls will be matched also on duration of membership at Kieser Australia, and all available fat free mass measurements available for control | We will utilise an interrupted time-series with control design and analysis using propensity score matching. This will be a standard regression model for this design including fixed effects of time, intervention, and period, time:intervention and period:time interaction, as well as period:intervention:time interaction. We will also include random intercepts by participant and utilise an autoregressive moving average (1) correlation structure. The primary estimand of interest is the contrast between the time coefficient (reflecting the main effect of RT over time) and the period:intervention:time interaction coefficient (reflecting the effect of GLP-1-RA over time). | Simulations were performed and parameterised to reflect prior knowledge from existing literature and knowledge of the Kieser Australia member population. We derived estimates for the effects of RT and GLP-1-RA on lean soft tissue mass from prior meta-analyses. We used the model of lean soft tissue mass loss during energy restriction and mitigation of this with concomitant RT to base our assumption of additive RT and GLP-1-RA effects. We also used previous meta-analysis of variance to examine whether interindividual response variation was present for either treatment and subsequently worked with the assumption of a fixed underlying effect. We also used previously reported measurement error for error terms in our | A statistically significant non-inferiority contrast of the time and period:intervention:time coefficients from our model will be interpreted as supporting our hypothesis that the effects of RT mitigate the lean soft tissue mass loss effect of GLP-1-RA treatment. A non-significant result will be interpreted as meaning we cannot conclude that the effects of RT are of a magnitude to mitigate the effects of GLP-1-RA treatment.<br>We will secondarily compare the magnitude of the effect estimates and confidence intervals to those reported in previous meta-analyses to qualitatively interpret whether the assumption of additive effects is supported i.e., where RT and GLP-1-RA | The theory that the effects of RT are potent enough to overcome the negative effects of GLP-1-RA upon lean soft tissue mass could be proven wrong by our results. Or more accurately given that we will test the non-inferiority of RT to GLP-1-RA when performed concomitantly, we could prove wrong the theory that RT is an insufficient intervention to mitigate the effects of GLP-1-RA treatment on lean soft tissue mass loss. |
|  | interrupted time-series analysis of propensity score matched individuals participating in RT and then either beginning GLP-1-RA treatment (intervention) or not (comparator). | and intervention groups extracted. |  | simulations for fat free mass from BIA.<br>We simulated 1000 datasets reflecting the Kieser Australia members of 24000 members using our empirical estimates of means, SDs, and correlations between variables, in addition to proportions of members with certain characteristics relevant to predicting GLP-1-RA propensity of treatment. We simulated propensity for GLP-1-RA treatment and checked this approximated estimates of treatment in the broader Australian population too. We also simulated missing data such that from the original 24000 member simulated datasets we were left with a sample size reflecting our estimates that ~6% of members have fat free mass measurements from BIA recorded.<br>We then used these 1000 datasets to simulate fat free<br>mass outcomes that were confounded by the variables used to determine propensity scores under an interrupted time-series design. We varied the number of measurements before and after treatment initiation, the autocorrelation coefficient magnitude, and also whether or not the effect of RT was of the magnitude reported in previous meta-analysis (i.e., additively exceeds over and above the GLP-1-RA effect), or was of a magnitude to only counteract it (i.e., the same magnitude as the GLP-1-RA effect but with an opposite sign such that it only prevented lean soft tissue mass loss).<br>We fit both models using inverse propensity of treatment weighting and also those using nearest propensity score matching.<br>Both revealed that we would likely achieve sufficient (i.e., >80%) statistical power to test the contrast of | effect estimates are similar to those reported where they have been used as single interventions this would support the assumption of additivity of effects. Also that the inference from fat free mass measurements by BIA are reflective of lean soft tissue mass changes. |  |
|  |  |  |  | RT and GLP-1-RA with the sample size projected (n=37 intervention, interquartile range= 8) irrespective of the modelling approach used. So, for pragmatic reasons we opted to use the matching approach which would reduce the burden of having to extract fat free mass measurements manually from clinical notes due to a smaller overall sample size (due to the smaller control group size). |  |  |
| **SECONDARY OUTCOME AND EXPLORATORY EFFECT ESTIMATION** |  |  |  |  |  |  |
| What is the effect of resistance training (RT) on strength during Glucagon-like Peptide-1 Receptor Agonist (GLP-1-RA) treatment? | Existing evidence is unclear as to the effects of GLP-1-RA treatment on strength, and as such we do not have any particular hypothesis regarding this. Thus, we will instead adopt an exploratory estimation approach. For strength outcomes, though secondary and exploratory, we will utilise the same approaches as above given the study design is the same. Whilst fat free mass measured by BIA is our primary outcome for which we have planned the study, we will also employ the same modelling for strength outcomes which, given the effects of RT on strength are relatively far larger in magnitude than its effects on lean soft tissue mass and we know already that we will have far greater strength outcome data for all Kieser Members as it is a core measurement taken at all review sessions, we anticipate also having sufficient power (in fact greater) as for our primary outcome and thus will yield relatively precise effect estimates. The only difference with the above analysis for fat free mass is that we will have multiple strength outcomes across exercises tested (e.g., chest press, leg press, pulldown, knee extension etc.). As such we will use a two-stage internal meta-analysis approach where for each outcome we will fit the same interrupted time-series with control model using propensity score matching, extract the contrast of interest estimate and standard error, and then synthesise these using a random effects meta-analysis to yield a single strength effect estimate. |  |  |  |  |  |

### Hypothesis

We hypothesize that RT will mitigate lean soft tissue mass loss, inferred through fat free mass measured by BIA, while adults are on GLP-1-RA treatment. Previous meta-analyses have provided estimates of both the magnitude of raw and standardised (i.e., intervention effect standardised for baseline standard deviation estimate; SMD) effects on lean soft tissue mass of RT (+1.61 kg, +0.27 SMD; (Benito et al., 2020)) and GLP-1-RA (−0.86 kg, −0.13 SMD; (Karakasis et al., 2025)) over similar intervention durations. Further, we assume additive effects of RT and GLP-1-RA treatment based on analogy with the energy restriction model of effects on lean soft tissue mass suggesting no interactive multiplicative effects between energy restriction and RT; for energy restriction alone the estimated SMD is −0.15 to −0.18 for a ∼100-200 kcal deficit and –0.24 to −0.26 for a ∼470 to 540 kcal deficit (Das et al., 2017), and for energy restriction with RT the estimated SMD is –0.31 for a ∼600 kcal deficit (Murphy & Koehler, 2022). Thus we specifically hypothesise that, in an interrupted time-series with control design of individuals participating in RT and then either beginning GLP-1-RA treatment (intervention) or not (comparator), the coefficient for the effect of RT over time (fixed effect estimate of time) will be non-inferior to the coefficient for the effect of GLP-1-RA over time (fixed effect estimate of period:intervention:time) i.e., a one-sided test of the contrast between these coefficients will show a statistically significant difference in favour of the RT coefficient, or synonymously the lower bound of the 90% confidence interval will be greater than zero.

## Methods

### Open science

All materials, code, and deidentified data are/will be available on the Github repository for this project (https://github.com/jamessteeleii/glp1_rt_cits) and linked to the corresponding Open Science Framework project page (https://osf.io/r4s9b/).

### Study design

This will be a retrospective study involving existing data from members of Kieser Australia. Kieser Australia is a private chain of franchised physiotherapy, exercise physiology, and resistance training clinics (at the time of writing with 30 clinics across Australia and ∼24000 members). Members participate in a standardised RT prescription of two sessions per week performing a selection of exercises on resistance machines to provide a full body workout (i.e., target all major muscle groups) in each session and are instructed to perform one set per exercise of repetitions to momentary failure. Members also typically engage in a review session approximately every three months in which they engage in both maximal isometric strength testing, review their progress, assess any existing or new clinical conditions, and conduct any additional measurements such as measurement of body composition. We have become aware through our exercise physiologists clinical notes and reporting that many members of Kieser Australia have begun GLP-1-RA treatment. As such, we identified this as an opportunity to utilise our existing data retrospectively to examine the effects of RT during GLP-1-RA treatment.

An interrupted time-series with control design (Lopez Bernal et al., 2018) will be employed wherein we will include a cohort who have, during their membership at Kieser Australia, begun participation in GLP-1-RA treatment (i.e., the intervention) alongside RT, and a group who have not begun GLP-1-RA treatment but have only participated in RT (i.e., the control). We will survey the membership of Kieser Australia via our internal surveying system to identify by self-report members who are currently, or have been previously, on GLP-1-RA treatment (i.e., the intervention) and the dates during which they were on these treatments. We will also confirm this, with consent from members, by having their exercise physiologist contact their GP to obtain their history of prescription. Members responding as having been on, or are currently on, GLP-1-RA treatment will then have their data queried in Kieser Australia’s database to identify those with body composition measurements. These are currently and historically recorded in clinical notes on our systems and not in a structured data format. As such, we will then manually identify from clinic notes information and dates of body composition measurements in order to extract fat free mass data from both before and during their treatment as fully as available. We will conduct propensity of treatment modelling (using logistic regression) of the entire Kieser Australia membership to derive direct propensity scores for our sample of individuals and then form a control group of members who have not reported having been on GLP-1-RA treatment and also have fat free mass measurements in clinical notes via nearest propensity score matching.

### Ethics approval

This project is deemed to fall under the exemption from a priori review and approval under the Australian NHMRC National Statement on Ethical Conduct in Human Research 2023, in addition to other guidelines such as those of the UK Research Integrity Office, as it considered to be low risk. The project will involve use of existing data only in deidentified form that is collected from Kieser Australia members with full informed consent already captured for research use as standard part of members contracts. The only additional prospective data capture will be the brief internal survey to members aiding us in identifying those from our membership who meet the characteristics for the intervention group; in this case, those who respond to self-report and identify themselves as having been on, or are currently undergoing, GLP-1-RA treatment. Internal member surveys are commonly conducted by Kieser Australia and do not require any a priori ethical approval under any existing legislative or guidance documentation that Kieser Australia is subject to.

### Blinding

Prospective blinding for this study is not possible. However, given the nature of the proposed retrospective study design we do not anticipate this introducing any significant bias. The individuals who collected the data, i.e. the physiotherapists, exercise physiologists, and exercise scientists within Kieser Australia clinics do so as a matter of business-as-usual service delivery and were unaware of the hypotheses of this study or prospectively that this study would even take place. Additionally, the analyses will be completed by the lead author of the project who has been independent from the collection of any data^2^ to reduce any possible bias and conducted under a strictly pre-specified analysis plan with clear estimands and hypotheses.

### Randomization

This is a quasi-experimental study design and so randomisation is not applicable for this study design.

### Participants

As noted, participants will be recruited from the current membership pool of individuals from Kieser Australia. The current membership of Kieser Australia at the time of writing based on demographic data in the Kieser Australia database are characterised as follows (mean ± standard deviation [SD]): 56.3±14 years old, 46% male (height 179±7.5 cm, weight 88.2±15.3 kg) and 54% female (height 165±7.3 cm, weight 73.6±17 kg), 42% have private health insurance, 0.4% have cardiovascular disease, 0.5% have type 2 diabetes, 15% have osteoarthritis.

A survey will be sent to all members at Kieser Australia via email and also a push-notification in their Kieser Konnect App (used to track their RT sessions), who will have the opportunity to complete the online survey within four weeks to assess their eligibility to be involved in the study and “opt in” to the study as part of the intervention group by self-identifying as having been on, or currently are on, GLP-1-RA treatment. A reminder will be sent two weeks after the original survey. Participants will thus include adults over the age of 18 years, who are or have been prescribed with GLP-1-RA treatment and have had body composition (including fat free mass) measured in a Kieser Australia clinic and recorded in their clinic notes. Participants will not be excluded from this study based on previous medical history but we will match based on propensity scores considering the demographic characteristics including those conditions noted above and described in the sections below. All participants will have also previously provided informed consent for their data to be used for research purposes in deidentified format as part of their Kieser Australia contract.

### Sample size

Sample size will be limited by resource constraints (Lakens, 2022); primarily the availability of data. Our simulations detailed below provide an estimated sample size, given existing data for Kieser Members, of median 37 [interquartile range: 8] participants in the intervention group. We will obtain a matched group of control participants of the same size as described below. Thus, the total sample size will be ∼74 participants in our estimation. This will provide us with sufficient statistical power as detailed below to test our hypothesis. If we identify a far larger sample of Kieser Australia members who are, or have been, on GLP-1-RA treatment allowing us to obtain a larger intervention sample size we will weigh up the benefit of this to generating mor precise effect estimates given resource constraints in the team as fat free mass data will have to be identified and manually extracted from member clinic notes. As such, a very large sample size would be prohibitively time-consuming to obtain data from our database in its current unstructured format.

### Rationale for sample size

Simulations were performed and parameterised to reflect background knowledge from existing literature and knowledge of the Kieser Australia member population. All code for the targets pipeline simulations is available in the online open materials. Firstly, we derived estimates for the effects of RT and GLP-1-RA on lean soft tissue mass from prior meta-analyses (Benito et al., 2020; Karakasis et al., 2025). A fixed effect –0.86 kg was used for the effects of GLP-1-RA treatment on lean soft tissue mass and a fixed effect of 1.6 kg for the effects of RT. These are approximately 12-week intervention effects and so, given time in our simulations was coded as week, were linearly transformed to reflect the slope for week. As noted above, we assumed additive effects which were justified based on the similar additive effects of energy restriction upon lean soft tissue mass as being analogous to the effect of GLP-1-RA treatment. We also assumed only fixed effects and no interindividual variability in effects (i.e., random effects) for either RT or GLP-1-RA. This is based on extensive evidence from large scale meta-analysis (Steele et al., 2023) that there is not inter-individual variation in the causal effects of RT on either strength or hypertrophy outcomes through comparison of variances in change scores between interventions and controls. Further, we extracted data from a previous meta-analysis of six placebo controlled trials of GLP-1-RA treatment and re-analysed this calculating both log transformed variance (lnVR) and coefficient of variation ratios (lnCVR) neither of which revealed evidence of inter-individual variation in the causal effects of GLP-1-RA treatment on lean soft tissue mass and if anything evidence against this in the case of lnCVR (lnVR = 0.02 [95% CI: −0.26 to 0.30; lnCVR; −1.33 [95% CI: −1.97 to –0.69]).

We then simulated 1000 datasets reflecting the Kieser Australia membership of 24000 members using our empirical estimates for characteristics, as noted above regarding participant population demographics, of means, SDs, and correlations between these variables (male age-height r = −0.188, age-weight r = −0.007, height-weight r = 0.399; female age-height r = −0.172, age-weight r = −0.002, height-weight r = 0.260), in addition to proportions of members with characteristics relevant to predicting GLP-1-RA propensity of treatment based on previous research (Radwan et al., 2025). We simulated propensity for GLP-1-RA treatment using the following odds ratios (age between 40-65 = 1.21; age over 65 = 0.38; has private health insurance = 0.81; is obese categorised by body mass index = 1.66; has cardiovascular disease = 0.75; has type 2 diabetes = 2.19; has osteoarthritis = 0.49) in an additive logistic model and then transformed to a propensity score on the probability scale and used the binomial distribution to simulate from this which simulated participants were in the intervention (i.e., undergoing GLP-1-RA treatment). This yielded approximately 2.5% of participants within simulated samples as being on GLP-1-RA treatment, which is not dissimilar to reported rates of prescribed use though may be conservative given off-label use rising (Kanellis et al., 2025; Lin et al., 2023; Radwan et al., 2025). Thus, we were content that our simulated propensity model was reasonably justified and produced a proportion of simulated members on the treatment that was realistic. Lastly, for each simulated dataset we also simulated missing data such that, from the original 24000 members simulated, datasets we were left with a sample size reflecting our estimates that ∼6% of members have fat free mass measurements recorded. This estimate of ∼6% of members having fat free mass measurements recorded is based on initial string matching in our database clinic notes for the following strings: “lean-mass”, “lean mass”, “lean-body mass”, “lean body mass”, “lbm”, “LBM”, “fat-free mass”, “fat free mass”, “ffm”, “FFM”, “muscle-mass”, and “muscle mass”.

We then used these 1000 datasets to simulate lean soft tissue mass outcomes in kilograms that were also influenced by the variables used to determine propensity scores under an interrupted time-series design. This introduced the confounding between treatment and our outcome of interest. The following fixed effect coefficients were used for the effects of confounding variables based on previous research where we were able to identify appropriate estimates, and using reasonable assumptions for those where we could not: the fixed intercept was based on the weighted mean of male and female means for estimated fat free mass (Kirk et al., 2021) reflecting the proportion of male and female members (which is also similar to the proportions of GLP-1-RA treatment users in the broader Australian population; (Lin et al., 2023)), and a random effect for the intercept was similarly weighted based on the SDs for males and females; the effects of age were set with fixed effects of 0.1 for those aged between 40 to 65 and –0.3 for those over 65 (Kirk et al., 2021); the fixed effect of height in centimetres was set at 0.15 (Baglietto et al., 2024); the fixed effect of weight in kilograms was set at 0.5 (Kulkarni et al., 2013); and finally the fixed effects of having private health insurance, cardiovascular disease, type 2 diabetes, and osteoarthritis were set at 1, −1, −1, and –1 respectively. Lastly, we set the residual error in the simulations based on approximate measurement error for fat free mass measured using bioelectrical impedance analysis at 2.5 (Vasold et al., 2019). We varied the number of measurements after baseline (k = 3, 4, 5, or 6) such that the total number of measurements was evenly distributed before and after treatment initiation and the total measurement was k+1, the autocorrelation coefficient magnitude (r = 0.1, 0.5, or 0.9), and also whether or not the effect of RT on lean soft tissue mass was of the magnitude reported in previous meta-analysis (+1.61 kg, i.e., additively exceeds over and above the GLP-1-RA effect), or was of a magnitude to only counteract it (+0.86 kg, i.e., the same magnitude as the GLP-1-RA effect but with an opposite sign such that it only prevented lean soft tissue mass loss).

To these simulated datasets we then fit two sets of models for interrupted time-series with controls: one using inverse propensity of treatment weighting and thus using all control members in the simulated dataset, and one using nearest propensity score matching and so using a sample size for the control group that was the same as the intervention. Both were standard regression models for interrupted time-series with controls designs including fixed effects of time, intervention, and period, time:intervention and period:time interaction, as well as period:intervention:time interaction^3^. They also both included random intercepts by participant and an autoregressive moving average (1) correlation structure. The primary estimand of interest extracted was the contrast between the time coefficient (reflecting the main effect of RT over time) and the period:intervention:time interaction coefficient (reflecting the effect of GLP-1-RA over time). For each simulation, this was extracted in addition to a non-inferiority test of the contrast i.e., a one-sided Wald test.

Across simulations the proportion of results with p values for this test that were below our type 1 error rate (alpha ≤0.05) were summarised with a binomial test^4^ and this reflected the estimated statistical power we would likely have for the study given all of our assumptions and estimated sample size. These revealed that even under the most extreme conditions of autocorrelation and number of measurements, we would likely achieve ∼80% statistical power as desired (see figure 1). Further, there was no discernable difference between the inverse propensity of treatment weighted or propensity score matching models. So, for pragmatic reasons we plan to use the propensity score matching approach which would reduce the burden of having to extract additional fat free mass measurements manually from clinical notes for the larger control sample that would be created with the inverse propensity of treatment weighted approach.

**Figure 1.**
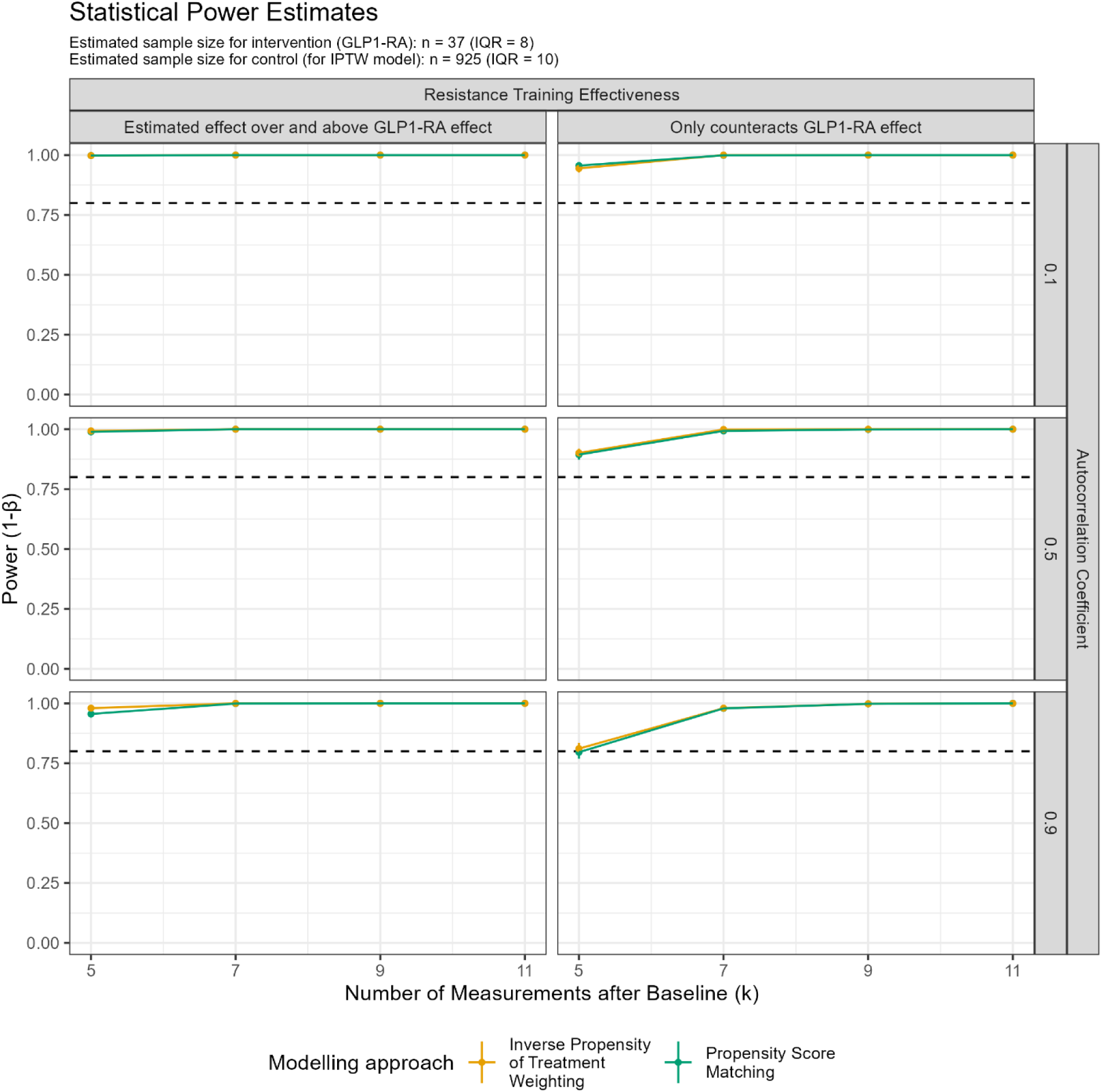
Estimates of statistical power from simulations (1000 simulated datasets per model and combination of conditions) varied across measurement number, fixed effect of resistance training, and autocorrelation coefficient. Note, IPTW = inverse propensity of treatment weighting, GLP-1-RA = Glucagon-like peptide-1 receptor agonist.

### Data collection procedures

As noted, participants will be identified from the current membership pool of Kieser Australia. A brief online survey, created using our internal surveying system, will be emailed to all participants (i.e. members at Kieser Australia), who will have the opportunity to be assessed for their eligibility to be involved in the study and “opt in” to the study as the intervention group. Note, survey logic will guide participants through relevant questions based on their preceding answers i.e., if they do not answer having been on any of the GLP-1-RA treatments then they will not answer corresponding questions that follow regarding them. The survey will include questions broadly related to if the member is on a GLP-1-RA for the purpose of intervention group identification, what type of GLP-1-RA, the intent for why the GLP-1-RA was prescribed, how long the member has been prescribed the GLP-1-RA, whether the member has been on more than one type of GLP-1-RA, and if the member has had any side effects since been prescribed the GLP-1RA. See Supplementary Table 1 to see the survey questions and answers. All data from this survey will be stored in the internal electronic database at Kieser Australia.

The internal electronic database will also have previously collected data that has been gathered for clinical reasons by a physiotherapist or accredited exercise physiologist, or as part of member review sessions including strength testing and patient reported outcomes, in addition to all member RT session data. Outcome data including body composition and strength results will be identified within the internal electronic database among the participants who have identified themselves as GLP-1-RA treatment users, in addition to matched control members.

Outcomes related to body composition, including weight, fat free mass (our primary outcome) and percentage of total body fat, will be manually extracted from clinic notes where they are recorded currently and historically within the internal electronic database held by Kieser Australia. Body composition will have been measured by an accredited exercise physiologist using a Tanita BC-545N bioimpedance scale following the standardized procedures recommended by the manufacturer as per Kieser Australia standard operating procedures. The time of when outcome data was measured will have been based on the clinical recommendations of the treating exercise physiologist and will likely differ between participants. All available measurements for members identified for the intervention of control groups will be extracted for use.

Secondary exploratory outcomes related to strength will have been assessed using maximal isometric strength tests on resistance machines that stabilise the joint and isolate the muscle during testing. Each test is typically conducted on a selection of exercise machines for each member depending on the specific nature of their training programme or needs. Peak torque is measured in Newtons, using a dynamometer placed in the shaft of the equipment. The tests are attempted three times with the highest strength measurement recorded following a standarsied protocol as per Kieser Australia standard operating procedures. Strength tests would have been conducted at the initial assessment by an exercise physiologist or physiotherapist (if deemed necessary) or at the fourth training session with an exercise scientist. Thereafter a strength test will be completed every ∼3 months at the members review session by an exercise scientist.

### Exercise intervention

Following the initial assessment with an exercise physiologist or physiotherapist, the participant will have completed a progressive RT program on resistance machines within the clinical facilities at Kieser Australia. Exercises completed by the participant will have targeted upper limbs, lower limbs, and trunk. One set will have been completed for each exercise at a 4-2-4 tempo (i.e. 4 second concentric contraction, 2 second isometric hold, 4 second eccentric contraction) until momentary failure was reached using a load that should permit this to occur in a range of 90 to 120 seconds^5^. Momentary failure was defined as members were unable to continue concentrically contracting and moving the load (Steele et al., 2017). If momentary failure was reached with a time-under-load exceeding 120 seconds, then load on the machine is increased. Members training independently will have been prompted to increase the load by ∼5% for the next session once the time-under-load was >120 seconds for the exercise. The training load for the next session was determined by an exercise scientist if the member was being supervised during the training session. Time-under-load of each machine has an upper limit of 180 seconds to ensure one Kieser member is not on the machine for too long and preventing other members from using that one machine in addition to discourage members from training with loads that are too light. Members are supervised 1:1 for the first six sessions (∼3-4 weeks) before transitioning into training independently or continuing to train supervised by an exercise scientist (note, in the subsequent matching process we will also ensure that this is a variable used in the propensity score model). A member’s program is also typically reviewed and may be altered in terms of exercise selection at each review session every ∼3 months.

### Data analysis

Statistical analysis of the data extracted will be performed in R, (v 4.4.3; R Core Team, https://www.r-project.org/) and RStudio (v 2023.06.1; Posit, https://posit.co/). All code and deidentified data will be available at the Github repository for this project (https://github.com/jamessteeleii/glp1_rt_cits) and linked to the corresponding Open Science Framework project page (https://osf.io/r4s9b/).

As noted, a standard model for interrupted time-series with controls (Lopez Bernal et al., 2018) using nearest propensity score matching for the control group will be utilised. Propensity score will be estimated using a Bayesian logistic regression model fit using the brms package and including the same fixed effects as those characteristics used in the propensity score model for simulations. Priors on the logit scale will be diffusely with a scale value of 10 but centered on the logit transformed estimates of the odds ratios used in the propensity score model for the simulation to reflect our prior knowledge of treatment propensity and improve precision of estimates of propensity scores.

The interrupted time-series with controls model will include fixed effects of time, intervention, and period, time:intervention and period:time interaction, as well as period:intervention:time interaction. It will also include random intercepts by participants and an autoregressive moving average (1) correlation structure. The model will be fit using the nlme package with the following syntax:

> lme(ffm ∼ time + intervention + time:intervention + period + period:time + period:intervention + period:intervention:time, random = ∼ 1 | participant, data = data, correlation = corARMA(form = ∼ 1 | participant, p = 1, q = 1))

Where ffm is the fat free mass measured by BIA, time is weeks (in this case weeks of participation in RT as a Kieser Australia member), intervention is a dummy coded variable which indicates if at a given point in time a participant is (1) or is not (0) on the intervention (i.e., GLP-1-RA), and period is similarly a dummy coded variable indicating whether it is the period before (0) or after (1) the introduction of the intervention. We will check for the presence of non-stationarity, autocorrelation, and seasonality effects as recommended for interrupted time-series analyses (Schaffer et al., 2021).

The primary estimand of interest will be the contrast between the time coefficient (reflecting the main effect of RT over time) and the period:intervention:time interaction coefficient (reflecting the effect of GLP-1-RA over time). This contrast will be tested using a non-inferiority test of the contrast i.e., a one-sided Wald test, using the marginaleffects package hypotheses() function. Alpha will be set at 0.05 as noted in our sample size estimation for power.

Whilst fat free mass is our primary outcome for which we have planned the study, we will also employ the same modelling for strength outcomes which, given the effects of RT on strength are relatively far larger in magnitude than its effects on lean soft tissue mass and we know already that we will have a far greater number of strength outcome data for all Kieser Members as it is a core measurement taken at all reviews, we anticipate also having sufficient p (in fact greater) as for our primary outcome and thus will yield relatively precise effect estimates. The only difference with the above analysis for fat free mass is that we will have multiple strength outcomes across exercises tested (e.g., chest press, leg press, pulldown, knee extension etc.). For strength outcomes, we will employ a similar approach as used in a recent randomised trial of RT interventions (Gschneidner et al., 2024) of z-scoring the strength outomes following the method of Penney (Penney, 2023) which yields z-scores that are comparable to a traditional Cohen’s d standardised mean effect. We will fit models to each exercise for which we have strength data and then conduct a two-stage internal meta-analysis by extracting the contrast estimates for the time and period:intervention:time coefficients and standard errors and then conducting a random-effects meta-analysis using the metafor package of these across exercises to yield a single test of the our hypothesis regarding the effects of RT on mitigating loss of strength during GLP-1-RA treatment. The justification for this approach is based on simulation for another recently pre-registered randomised trial the lead author has conducted which showed that, under conditions of heterogeneity of variances when assessing different operationalisations of the same underlying construct (i.e., “strength” in this case), the two-stage approach offers greater statistical power due to more appropriate weighting of the estimates (see registration at https://osf.io/bxa8p/).

### Data cleaning procedure

Data will be cleaned before any analyses are performed because of the potential size of the internal database and that outcome data and data regarding the exercise intervention can be entered in manually by the staff at Kieser Australia. Participants (Kieser members) also can manually enter data related to the exercise intervention if they are training independently. Thus, impossible values for exercise loads will be filtered based on known minimum and maximum loads on the exercise machines utilised. This is so that we can report descriptively the characteristics of the RT intervention completed (not just prescribed). We will also exclude any members data from our analysis where they have not completed a minimum of 80% of the prescribed RT program (i.e., twice a week) over the duration of observation included and so also match on this per-protocol aspect of the comparator.

Data related to the outcomes of interest or the exercise intervention that are outside of plausible ranges (with respect to its unit of measurement) are likely due to error in data entry. We adopted this method of cleaning which has similarly been used for outcome measures in other large registry database preparation (Steele et al., 2021). Plausible ranges for data (outside of the survey in Supplementary Table 1) can be viewed in Supplementary Table 2. If data has been entered incorrectly, i.e. outside of the plausible ranges, then it will be excluded from the analyses, as it would not be possible to address the possible reason for the incorrect input.

### Registered Report for existing data

We believe that this proposed Registered Report achieves level 4 i.e., *“At least some data/evidence that will be used to answer the research question already exists AND is accessible in principle to the authors (e.g., residing in a public database of with a colleague) BUT the authors certify that they have not yet accessed any part of that data/evidence”.* This is because, whilst all of the outcome data are available to us currently, we do not have access to data specifically pertaining to this research question given that we do not know systematically which members of Kieser Australia are currently, or have previously been, on GLP-1-RA treatment as a research team. Clinicians at Kieser Australia may or may not have knowledge of this and indeed may have included it in their clinic note for which we do have access. However, we attest that as a research team we have not undertaken any searches of this text data to identify any members.

## Results & Discussion

Results and discussion will be included upon completion of the study in a Stage 2 report.

## Data Availability

https://osf.io/r4s9b/

## Contributions

All authors conceived the idea for the study. JS and MNM designed the study, will provide data curation, project administration, and wrote the initial draft of this stage 1 manuscript. Simulations and formal analyses were conducted by JS. All authors contributed to editing and approving this stage 1 manuscript.

---

**Supplementary Table 1.**
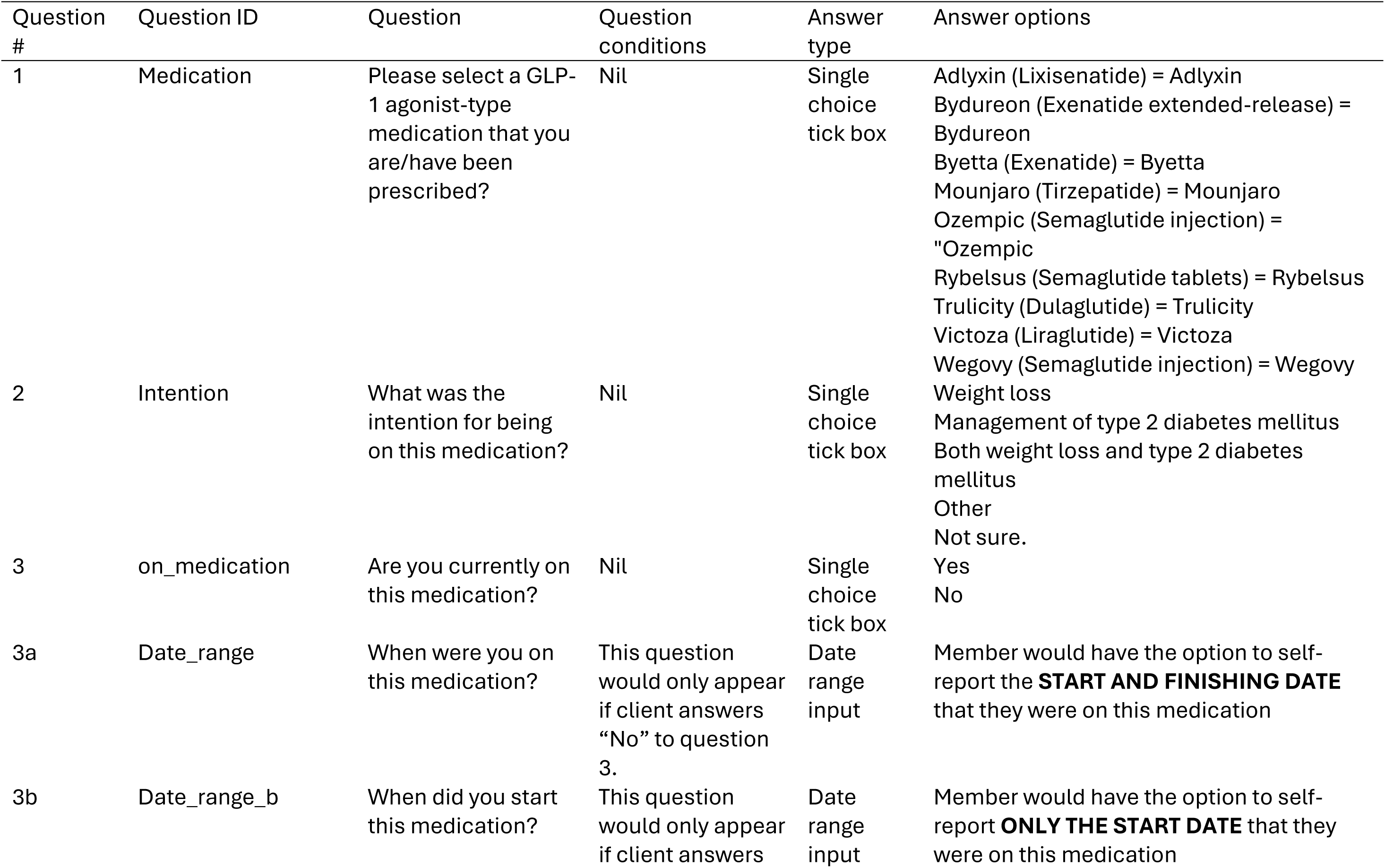
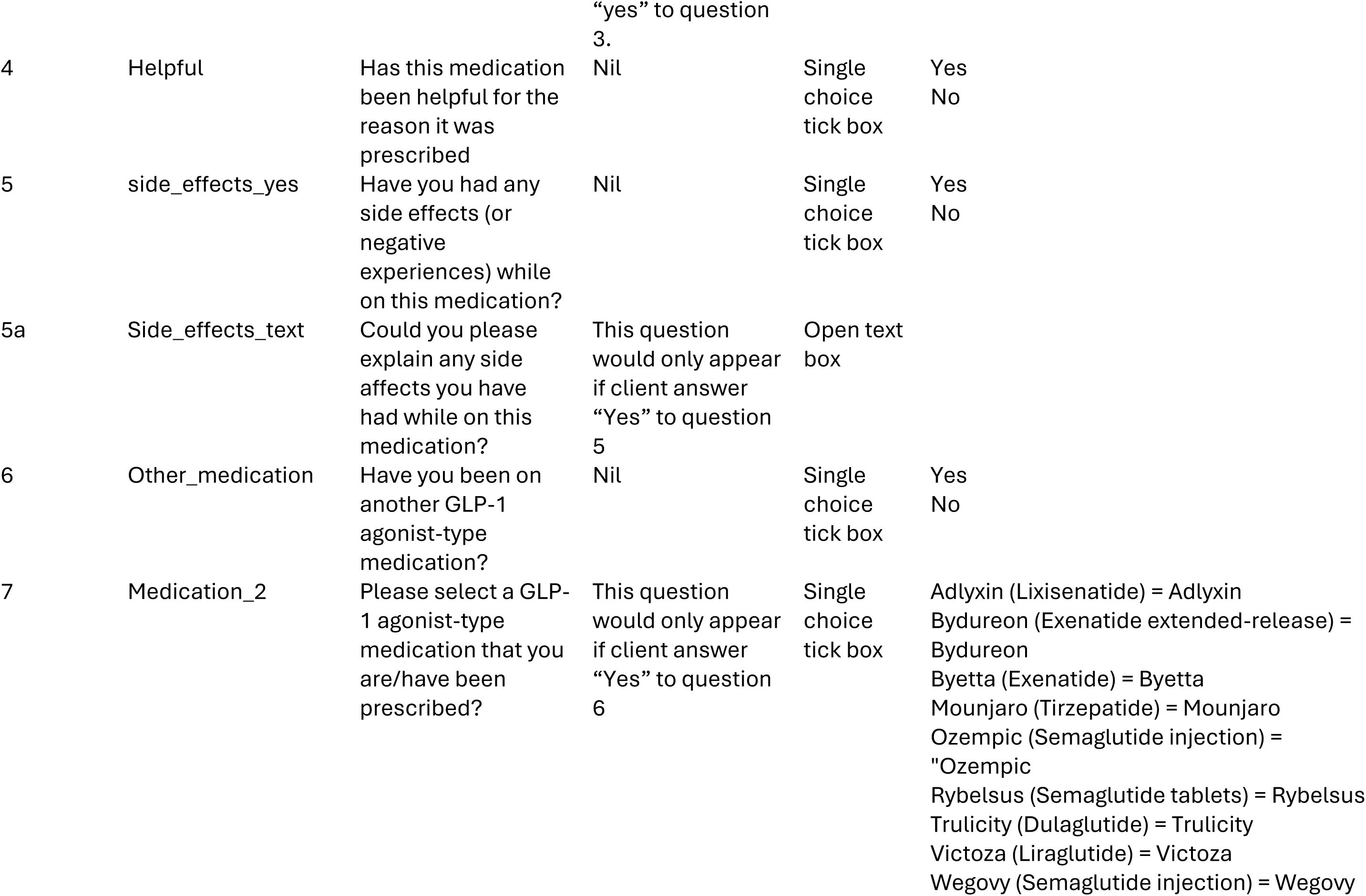
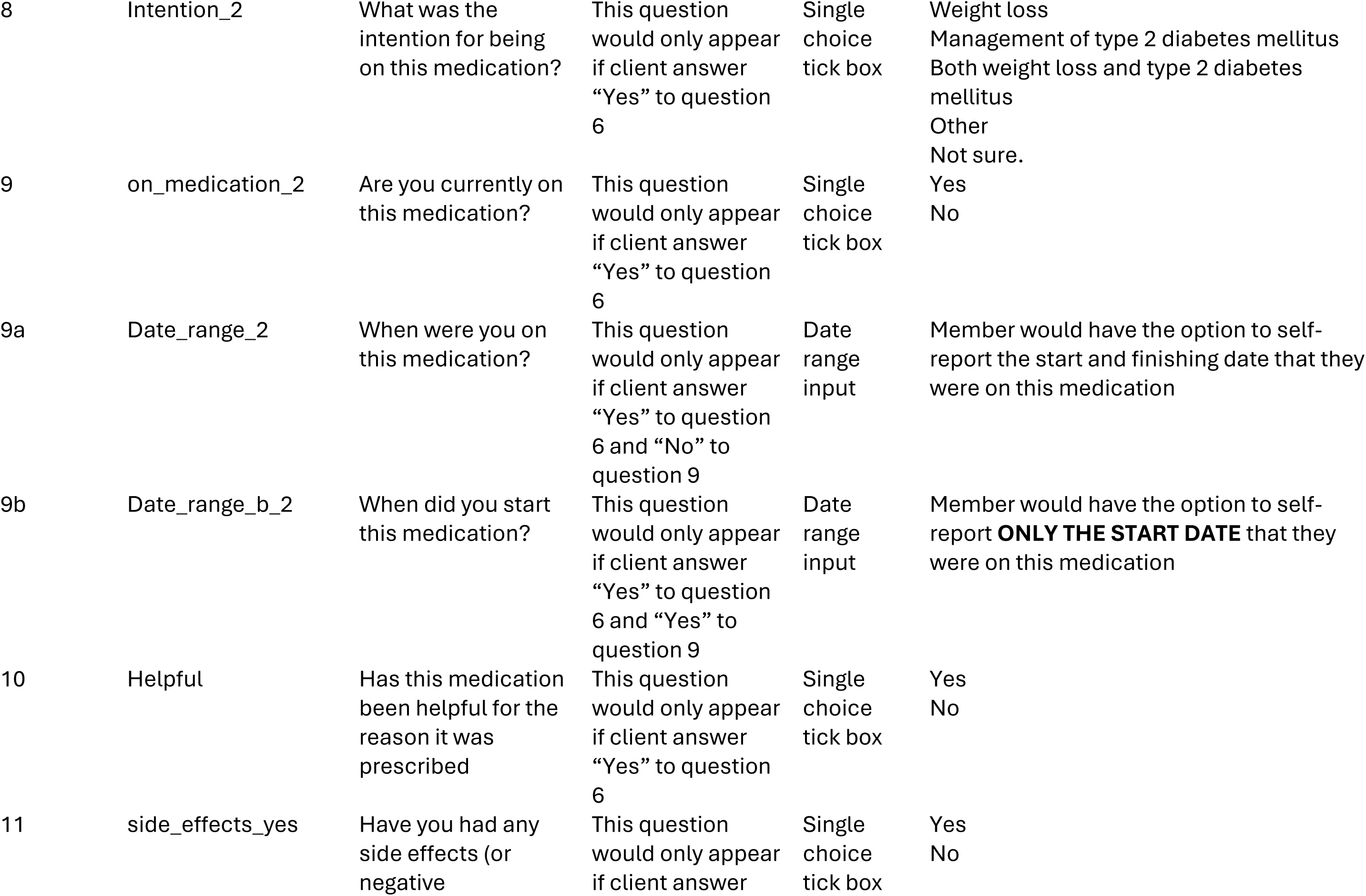
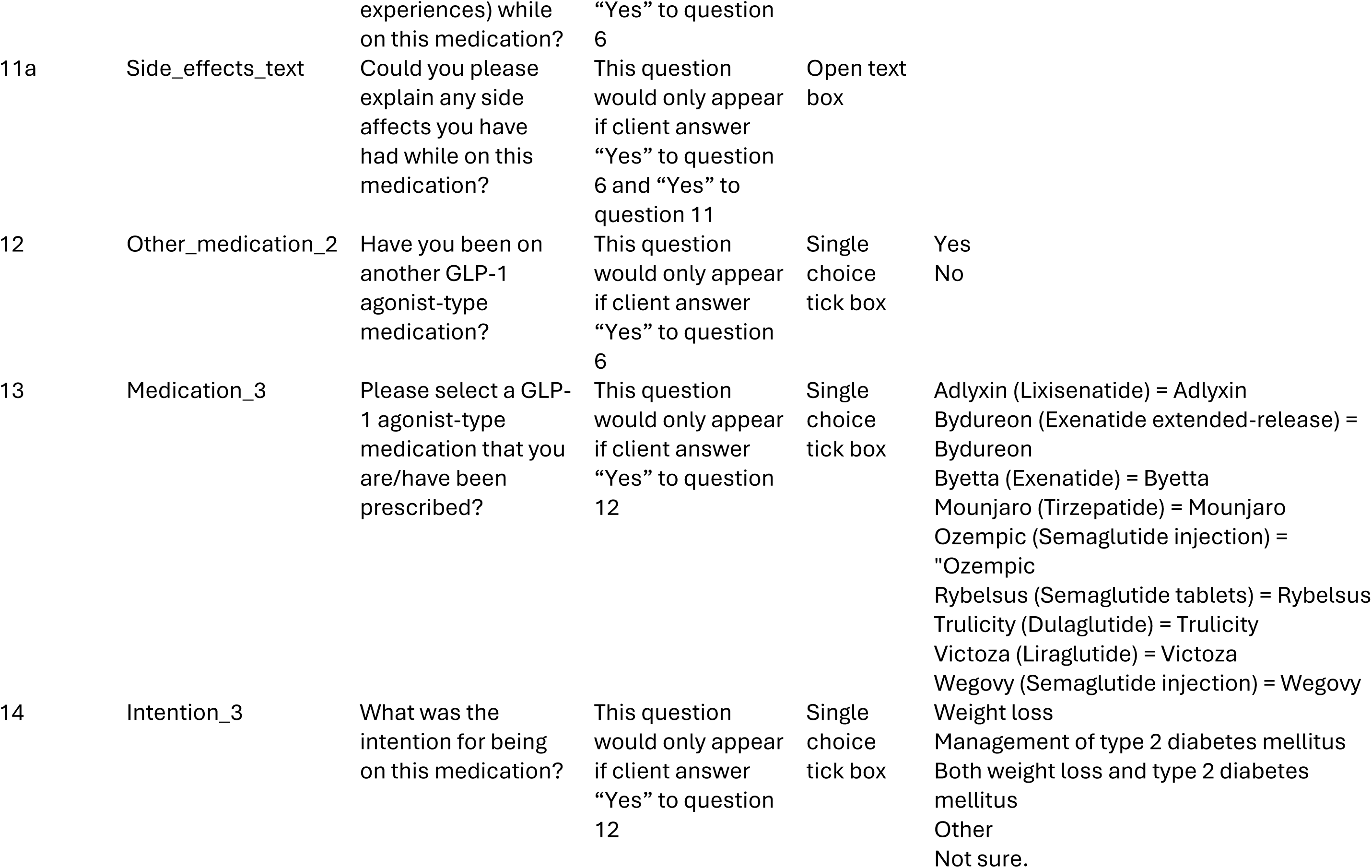
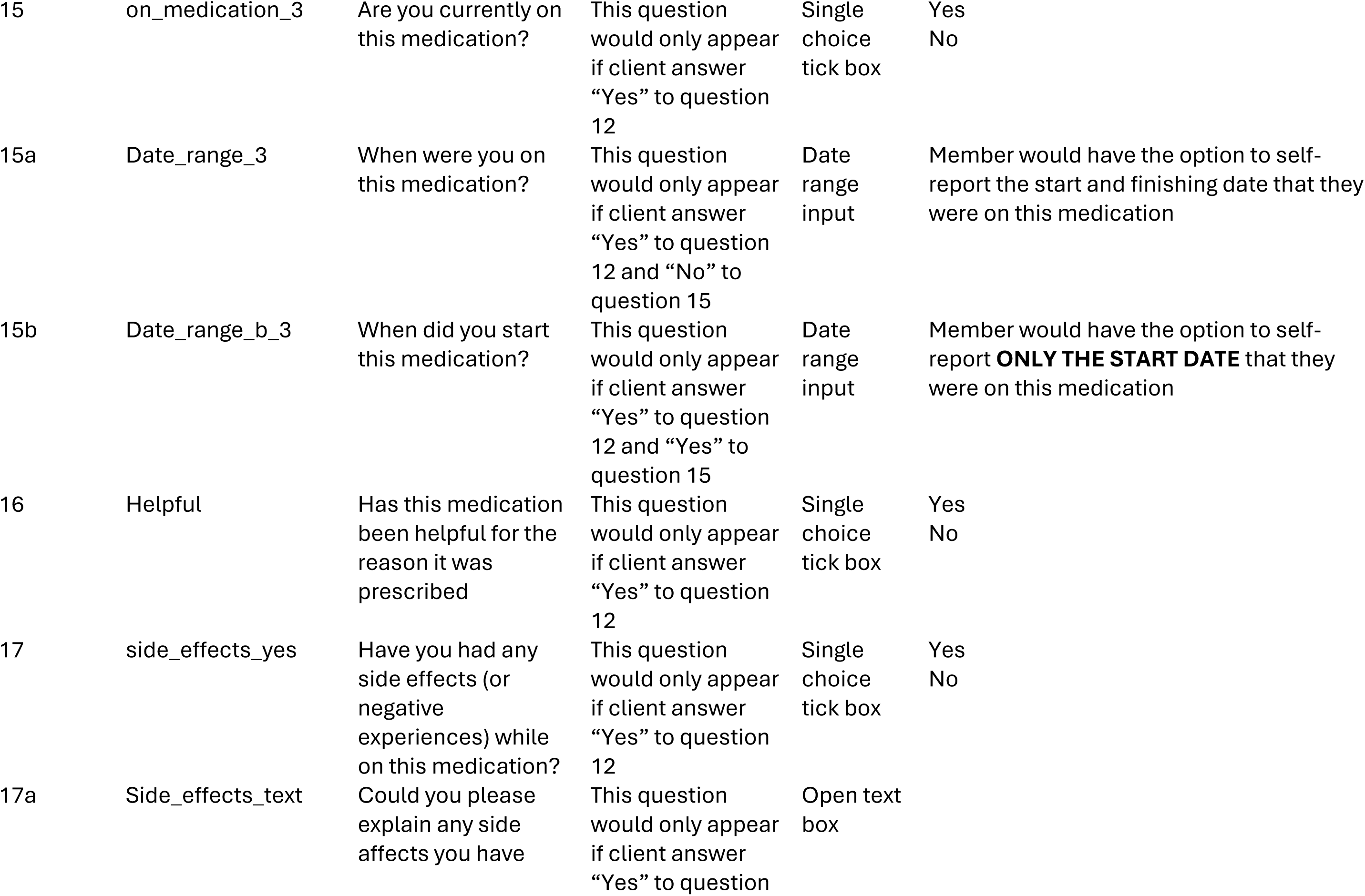
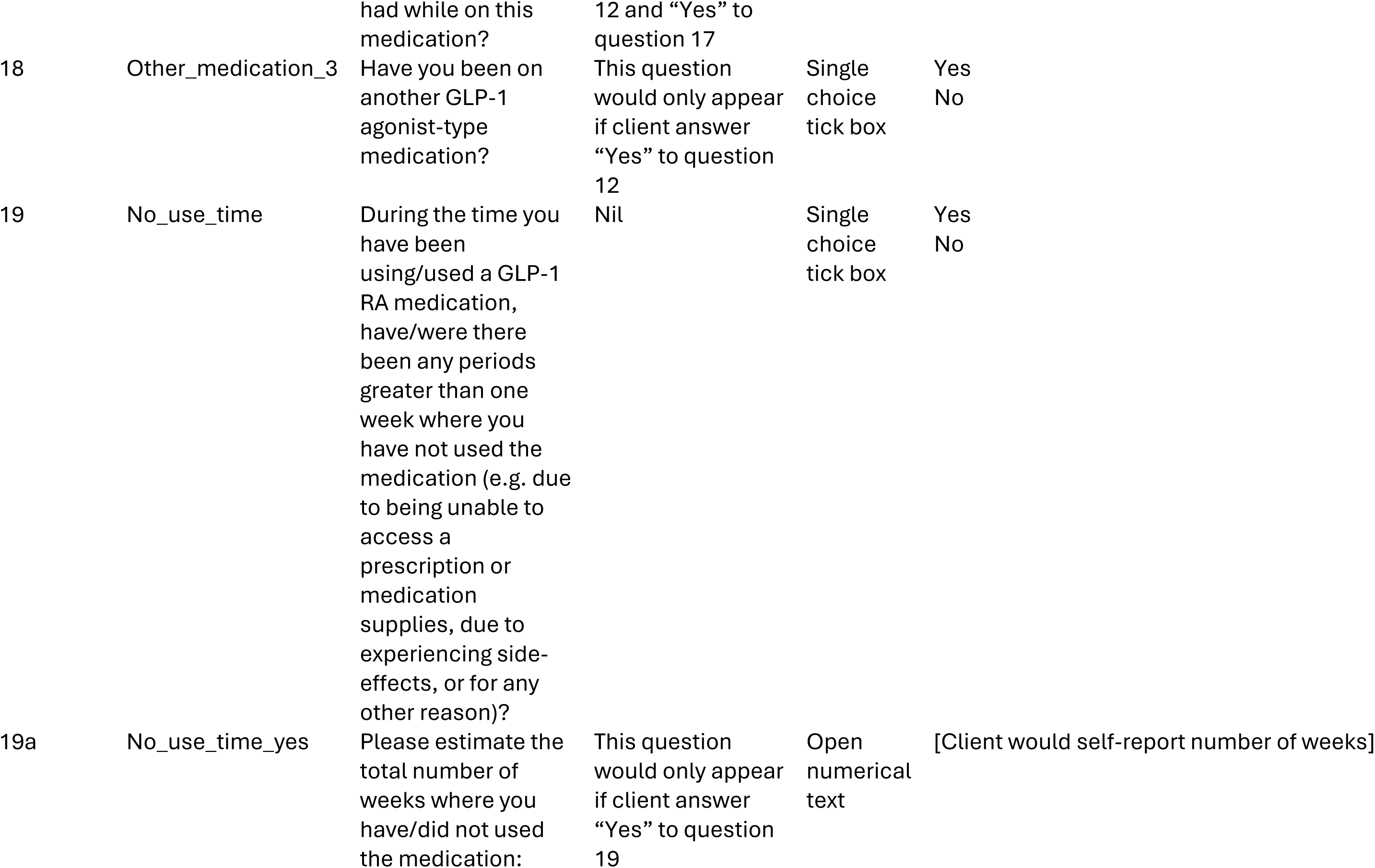
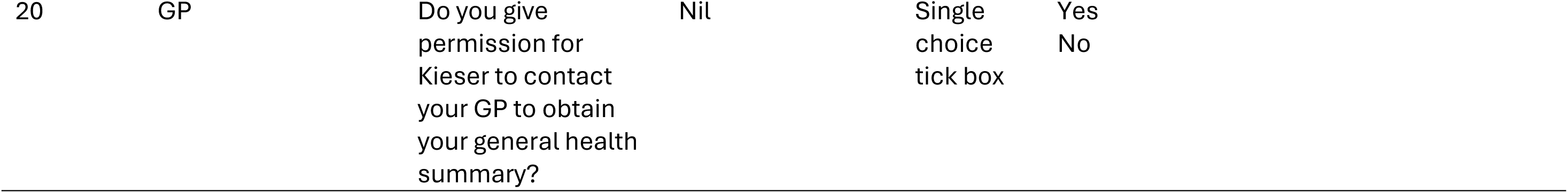
Survey questions and answers to identify members at Kieser Australia that are one GLP-1Ras.

**Supplementary Table 2.** Ranges used for data cleaning.

| Variable | Measurement and units | Range |
| --- | --- | --- |
| Age | Years | Cleaned to range of 0 to 100 |
| Height | Centimeters (cm) | Cleaned to range of 122.5 cm to 205 cm |
| Weight | Kilograms (kg) | Cleaned to range of 40 kg to 150 kg |
| Body mass index | Kilograms Per Metre Squared (kg.m<sup>2</sup>) | Cleaned to range of 12 kg.m<sup>2</sup> to 75 kg.m<sup>2</sup> |
| Body fat percentage | Percentage (%) | Cleaned to range of 5 – 70 % |
| Fat-free muscle mass | Kilograms (kg) | Cleaned to range of 18.7kg to 100kgs |
| Time under load | Seconds | Cleaned to range of 0 seconds to 180 seconds |
| Exercise weight | lbs | Cleaned to range of 0 lbs to 498 lbs |
| Strength test | Newtons | Values exceeding 3 standard deviations of baseline strength test data in Kieser Australia database |

## Footnotes

1 Note, strictly speaking the fat free mass that is measured through bioelectrical impedance analysis (BIA) is not limited to lean soft tissue mass (Heymsfield et al., 2024) and it should not necessarily be assumed that fat free mass loss from large weight loss such as through GLP-1-RA treatment is due to lean soft tissue loss or skeletal muscle loss (Tinsley & Heymsfield, 2024). However, given the use of RT as our comparator which mostly affects lean soft tissue and skeletal muscle mass, and the pragmatic reason that fat free mass measured by BIA is what we will have access to, we feel that it is justified to interpret fat free mass as indicative of lean soft tissue mass in the present study. Indeed, as we note in table 1, we will compare estimates of the effects of RT in our present study to those reported for lean soft tissue mass in previous large meta-analysis to check this assumption.

2 They are based remotely in the UK primarily.

3 The supplementary materials from (Lopez Bernal et al., 2018) provide a visualisation and explanation of what each of the fixed effect coefficients in this model parameterisation refer to in the context of an interrupted time series with control design for reference (link <u>here</u>).

4 Note, the number of trials in the binomial test was adjusted to reflect the total number of simulations without error as in some cases of simulation models may not converge. In the present simulations for the inverse propensity of treatment weighted models only 6 of the 24,000 models failed, and for the propensity score matching 411 of the 24,000 models across all conditions.

5 Notably, we are aware that many members do not train to momentary failure despite being prescribed to do so whether they are supervised or not, but we do still see meaningful strength changes in our members and so assume the intervention is efficacious - see (Steele et al., 2025)

